# A Cohort Study: Evaluating Self-Efficacy in Adolescents Attending a Tailored Youth-Informed Breastfeeding Program

**DOI:** 10.1101/2020.02.10.20021808

**Authors:** Christina M. Cantin, Wendy E. Peterson, Amisha Agarwal, Jemila S. Hamid, Bianca Stortini, Nathalie Fleming

## Abstract

Adolescents (≤19 years of age) have lower rates of breastfeeding (BF) compared to older mothers. BF self-efficacy (SE), defined as a mother’s confidence in her ability to breastfeed her infant, has been identified as an important factor influencing BF outcomes. An innovative youth-informed BF program for young women was designed and implemented, which included staff training, a prenatal BF class and BF peer support. The objective of this cohort study was to evaluate the effectiveness of the program in improving young mother’s BF SE.

Participants were pregnant adolescents recruited from a large urban non-profit social service outreach centre. The Breastfeeding Self-Efficacy Scale-Short Form (BSES-SF) was administered to participants before and after participating in the BF program. BSES-SF scores were summed to determine a composite score and compared descriptively using mean score. Un-aggregated, item-by-item, comparison of pre-vs post-BF program scores were also compared to examine improvements in SE. A total of 20 adolescent mothers (mean age = 16.6) attended the BF program. An increase in the total BSES-SF score was observed based on descriptively comparing the mean pre vs post intervention.

Prenatal education and peer support adapted to the needs of adolescent mothers was associated with increased BSES-SF. These results are promising given that clients attending programs at this agency have low-income, low educational attainment, variable family support, housing instability, and are at-risk for not breastfeeding. Future studies with larger cohort are required to further validate and establish generalizability, as well as to determine the effect on BF duration rates.

## Introduction

Adolescents, defined as women less than 20 years of age, have a lower rate of breastfeeding (BF) and traditional BF support given to older mothers may not meet the needs of pregnant and parenting youth (Best Start Resource Centre, 2015a; Sipsma, Jones, & Cole-Lewis, 2015; Sipsma, Magriples, Divney, Gordon, Gabzdyl, & Kershaw, 2013). Given that the benefits of BF are well known, the World Health Organization recommends exclusive BF up to 6 months of age, with continued BF along with complementary foods up to 2 years of age or beyond (World Health Organization, 2020). The benefits of BF can be even more significant for the young mother. As a large proportion of young mothers come from a low socio-economic environment, the financial and health savings can be important advantages as well (Sipsma et al., 2015). Other BF benefits include increased intrapartum interval (lactational amenorrhea), decreased incidence of infant infections, enhanced maternal-infant bonding, and reduced risk of postpartum depression. Finally, because of the higher rates of postpartum depression and violence in young mothers, BF can enhance mother-infant bonding and thus be a cornerstone for young mothers to help create safer and loving families (Sipsma et al., 2015).

BF initiation rates among Canadian women have increased significantly in the last decades. In 1965, less than 25% of mothers breastfed compared to 89% in 2011, however, only 26% of the women who initiated BF in 2011-2012 continued to breastfeed their infants exclusively for 6 months (Gionet, 2013; Health Canada, 2015). Public health reports and studies have identified that adolescent mothers (15-19 years) have lower rates of BF initiation, BF at time of hospital discharge and BF continuation compared to adult women (Best Start Resource Centre, 2015a; Sipsma et al., 2015; Sipsma et al., 2013). Higher rates of initiation and continuation have been reported when young women received BF education and counselling (Durham Region Health Department, 2015; Fleming, Ng, Osborne, Biederman, Ill, Dy, … Walker, 2013; Fleming, Tu, & Black, 2012; Sipsma et al., 2013).

The majority of existing interventions to increase BF rates target adult women. The studies show that education and support given by laypeople or professionals is effective in increasing BF duration (Sipsma et al., 2015). Compared to adults, Ontario adolescents have a 27% lower rate of exclusive BF at hospital discharge (Fleming et al., 2013). Furthermore, a large retrospective population-based cohort study in Ontario, consisting of 22,023 adolescent women less than 20 years of age, revealed that only half (48.8%) *exclusively* breastfed their infant at time of hospital discharge (Leclair, Robert, Sprague, & Fleming, 2015). Nevertheless, none of the existing interventions focus on adolescent mothers.

Adolescent mothers encounter multiple challenges which contribute to the low rates of intention, initiation and duration of BF. These challenges include: limited access to the social determinants of health (poverty, food and housing insecurity); negative effect on maternal confidence and attitudes by lower educational levels, low BF intention/confidence, possible negative attitude towards BF; high risk behaviours in pregnancy (smoking and substance use); higher rates of mood disorders and violence in pregnancy; lack of accessible support and adolescent friendly services (Apostolakis-Kyrus, Valentine, & DeFranco, 2013; Nesbitt, Campbell, Jack, Robinson, Piehl, & Bogdan, 2012; Sipsma et al 2013). In a retrospective population based-cohort study, Apostolakis-Kyrus and colleagues found that the most significant factors contributing to a lack of BF initiation in adolescent mothers was unmarried status and socioeconomic disadvantage.^11^

Adolescents are also faced with simultaneously experiencing issues of adolescence and motherhood: integrating new parental roles and romantic relationships, struggles with self-esteem and self-image and loss of former peer groups (Sipsma et al., 2015). Hence, the interventions to increase BF rates in young mothers need to be tailored to this population in order to yield positive outcomes (Leclair et al., 2015; Sipsma et al., 2013). Indeed, tailored pre-and postnatal care may have a pivotal role to improve BF education, intention, initiation and duration in this population (Fleming et al., 2012; 2013; Sipsma, 2013). BF peer support has been identified as a promising strategy to reach and support populations, such as young mothers, with lower rates of BF (Best Start Resource Centre, 2015a; 2015b). Peer support provides a low cost, non-medical intervention by caring, passionate and non-judgmental women with BF experience (Best Start Resource Centre, 2015c).

Self-efficacy has been found to be an important factor associated with BF duration and exclusivity. Studies of adolescent mothers demonstrate that BF self-efficacy (SE), both pre- and postnatally, is an important factor influencing BF outcomes (Dennis, Heaman, & Mossman, 2011; Mossman, Heaman, Dennis, & Morris, 2008). BF SE is defined as the mother’s confidence in her ability to breastfeed her infant (Dennis et al., 2011). Adolescents with low post-natal BF SE are nearly 4 times more likely to quit BF prior to 28 days postpartum (Mossman et al., 2008).

Informed by these research findings and guided by our own experience working with adolescents (Ontario Ministry of Health and Long Term Care, 2014), we designed and implemented an innovative youth-informed BF program in 2015 (Best Start Resource Centre, 2017), which combines three evidence-based interventions to support young women and improve their BF outcomes: staff training in BF best practices, a revision of prenatal BF curriculum to be youth-friendly, and a new BF peer support group.

The objective of this research was to conduct a cohort study to evaluate the effectiveness of the tailored BF program in improving BF SE. Our hypothesis was that adolescent mothers would demonstrate increased BF SE after attending one prenatal BF class and /or four BF peer support sessions.

## Methods

### Design

A prospective pre and post cohort design was used, and a survey was administered before and after the intervention. Ethical approval was obtained from the University of Ottawa Research Ethics Board (#H02-15-09B) for this cohort study.

### Setting

The study was conducted in a Canadian non-profit social service agency that provides a full range of services for young pregnant women, young mothers and fathers (<25 years), and their children (<3 years of age). This agency supports young women to have a healthy pregnancy and birth, and helps young parents gain the knowledge and skills to raise healthy and happy children. There is a residence for up to 10 pregnant and five postpartum clients and their babies, as well as an outreach centre which offers a one-stop location for health and social programming and services.^19^ Women attending programs at this agency in general have low-income, low educational attainment, variable family support, and housing instability. Consequently, the majority of clients receives social assistance and relies on supports such as the food bank and the clothing bin.

### Participants

The targeted study participants were adolescents (19 years of age or less) currently registered as clients at the agency described above. The following inclusion criteria was used 1) currently pregnant or parenting, 2) attending a prenatal BF class or the BF peer support program, and 3) could read and write in English. Participants older than 19 years of age were excluded from our analysis. Participants were recruited at the beginning of the prenatal BF class or at beginning of the first BF peer support session they attended. Recruitment was done between November 2015 and October 2016 by verbal invitation by agency staff at selected programs, and by posting flyers in the common areas of the agency. Participants were eligible to participate in the study if they met the inclusion criteria.

### Intervention

In 2015, we developed the *Youth-Informed Breastfeeding Program* to address a gap in BF support. This program included staff training, a revision of prenatal BF class curriculum and the creation of a new BF peer support program. The staff training consisted of a two-hour session focused on evidence-based BF support for young mothers. A public health nurse, who was an International Board Certified Lactation Consultant (IBCLC), provided this training which included an oral presentation, interactive discussion and case studies. This was offered on two separate days to allow all staff at the agency to participate in the training, regardless of their role (e.g. receptionist, administrators, program coordinators, residence staff etc.).

The prenatal BF class was one session in a series of eight prenatal classes on various topics (the name of the program is *Pregnancy Circle*). The 2-hour class was facilitated by a public health nurse from a local public health agency, which received the Baby-Friendly Initiative Designation in May 2013 and thus the curriculum was consistent with the Baby Friendly Initiative Standards.^20, 21^ This class was available to pregnant youth and their partners or support persons. The curriculum was revised to reflect the realities of young mothers such as their single status, their attendance at school versus employment, and common social situations that would be relevant to youth.

The weekly BF peer support sessions (1.5 hours) were co-facilitated by a program coordinator from the agency where the study was being conducted and a youth leader who received peer-mom leader training (the name of the program is *Little Milk Miracles*). There were four sessions that covered a range of topics under the following themes: getting off to a good start, BF in the early days, troubleshooting common concerns, and continuing to BF. Childcare, transportation and a healthy snack were provided as this facilitated young mothers’ participation in the program. Prenatal or postnatal youth could attend the peer support sessions.

Peer-mom leader training consisted of four hours of training which was delivered over two sessions by the program coordinator and a public health nurse, who was an IBCLC. The training consisted of interactive discussions and case studies which enabled participants to review basic BF information and to better understand the role and expectations of peer-mom leaders as well as learn about available community resources. Peer-mom leaders were selected based on having 4 months or more breastfeeding experience, possessing qualities conducive to the role (i.e. approachable, positive, supportive), and a satisfactory police record check.

### Measurements

To measure BF SE, we developed a pre and post questionnaire. The pre-questionnaire included twelve demographic questions: age, marital status, employment status, obstetrical history, parenting status, current pregnancy status (if pregnant, # of weeks), BF education received, previous BF experience (yes/no, if yes for what duration), concerns related to BF, awareness/knowledge of community resources, how they heard about the program, and reason they came to the program.

The Breastfeeding Self-Efficacy Short Form scale (BSES-SF) was used in both the pre and post questionnaire as it has been validated for use with adolescent mothers (Dennis et al., 2011; Mossman et al., 2008). The BSES-SF is a 14-item self-report instrument measuring BF confidence. Each item is a 5-point Likert-type scale question where 1 indicates “not at all confident” and 5 indicates “always confident”.

### Data Collection

Data collection occurred between November 2015 and October 2016. Participants who consented to participate in the study were recruited from the Pregnancy Circle prenatal classes and/or the Little Milk Miracles peer-support program. *Prenatal BF class*: Participants completed the pre-intervention questionnaire prior to attending the BF class and they completed the post-intervention questionnaire at the end of the two-hour class. *BF Peer-support*: Participants completed the pre-interventions questionnaire at the beginning of the first BF peer support session they attended and they completed the post-intervention questionnaire after attending four sessions. Participants were given approximately 15 minutes to complete the paper-based surveys.

### Statistical Analysis

Demographic variables for the study participants were summarized using descriptive statistics, where mean and standard deviation (SD) or median and interquartile range (IQR) were used for continuous outcomes. Frequency and percentages were used for categorical variables. Important concerns about breastfeeding and reasons for attending the program were summarized in a similar manner. For all participants with complete data (i.e. all questionnaire items answered pre and post), total scores were aggregated by summing values across all questions and mean composite scores were calculated for both pre- and post-intervention data. This was done at an overall level with data from both the prenatal and peer-support group combined; we also performed analysis for each intervention group (prenatal class and peer-support group) separately. Comparisons between pre- and post-intervention measurements were made descriptively, by comparing mean and median values between the groups. Pre- and post-intervention scores for each question was also analyzed descriptively and visually to evaluate improvements in selected items related to young mothers’ BF confidence after attending the BF program. All statistical analyses were conducted in the R statistical software, version 3.4.1 (R Core Team, 2017).

## Results

Full demographic data are provided in Table 1. In total, 20 adolescents 19 years of age and under participated in the study, of whom 90% (N = 18) said they were pregnant. The mean age of participants was 16.6 (SD= 1.3) years and most reported that they had a partner/boyfriend (N=12; 60%). The majority of the participants had never been pregnant before (n=11; 57.9%) and therefore few had BF experience (n=2; 10.5%).

**Table 1.**
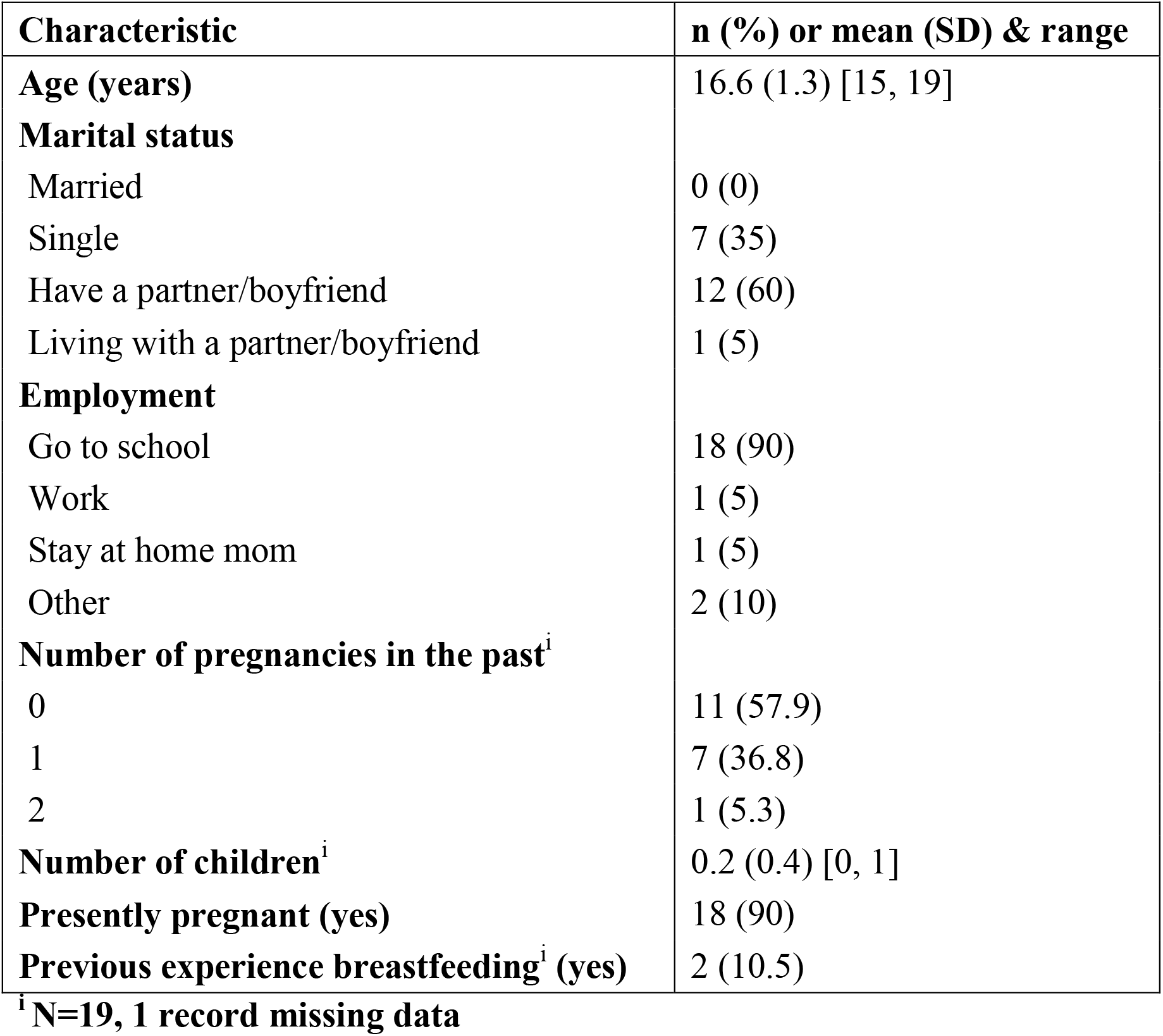
Demographics (N=20)

| Characteristic | n (%) or mean (SD) & range |
| --- | --- |
| **Age (years)** | 16.6 (1.3) [15, 19] |
| **Marital status** |  |
| Married | 0 (0) |
| Single | 7 (35) |
| Have a partner/boyfriend | 12 (60) |
| Living with a partner/boyfriend | 1 (5) |
| **Employment** |  |
| Go to school | 18 (90) |
| Work | 1 (5) |
| Stay at home mom | 1 (5) |
| Other | 2 (10) |
| **Number of pregnancies in the past<sup>i</sup>** |  |
| 0 | 11 (57.9) |
| 1 | 7 (36.8) |
| 2 | 1 (5.3) |
| **Number of children<sup>i</sup>** | 0.2 (0.4) [0, 1] |
| **Presently pregnant (yes)** | 18 (90) |
| **Previous experience breastfeeding<sup>i</sup> (yes)** | 2 (10.5) |

<sup>i</sup>N=19, 1 record missing data

The participants identified a variety of concerns (Table 2) which included: BF in public (n=7; 35%), being unable to start BF (n=6; 30%), not knowing what is normal with respect to BF (n=6; 30%), and not having enough information about BF (n=6; 30%). Not surprisingly, the majority of participants identified wanting to learn more about BF (n=17; 85%) as their main reason for attending the program (Table 2).

**Table 2.**
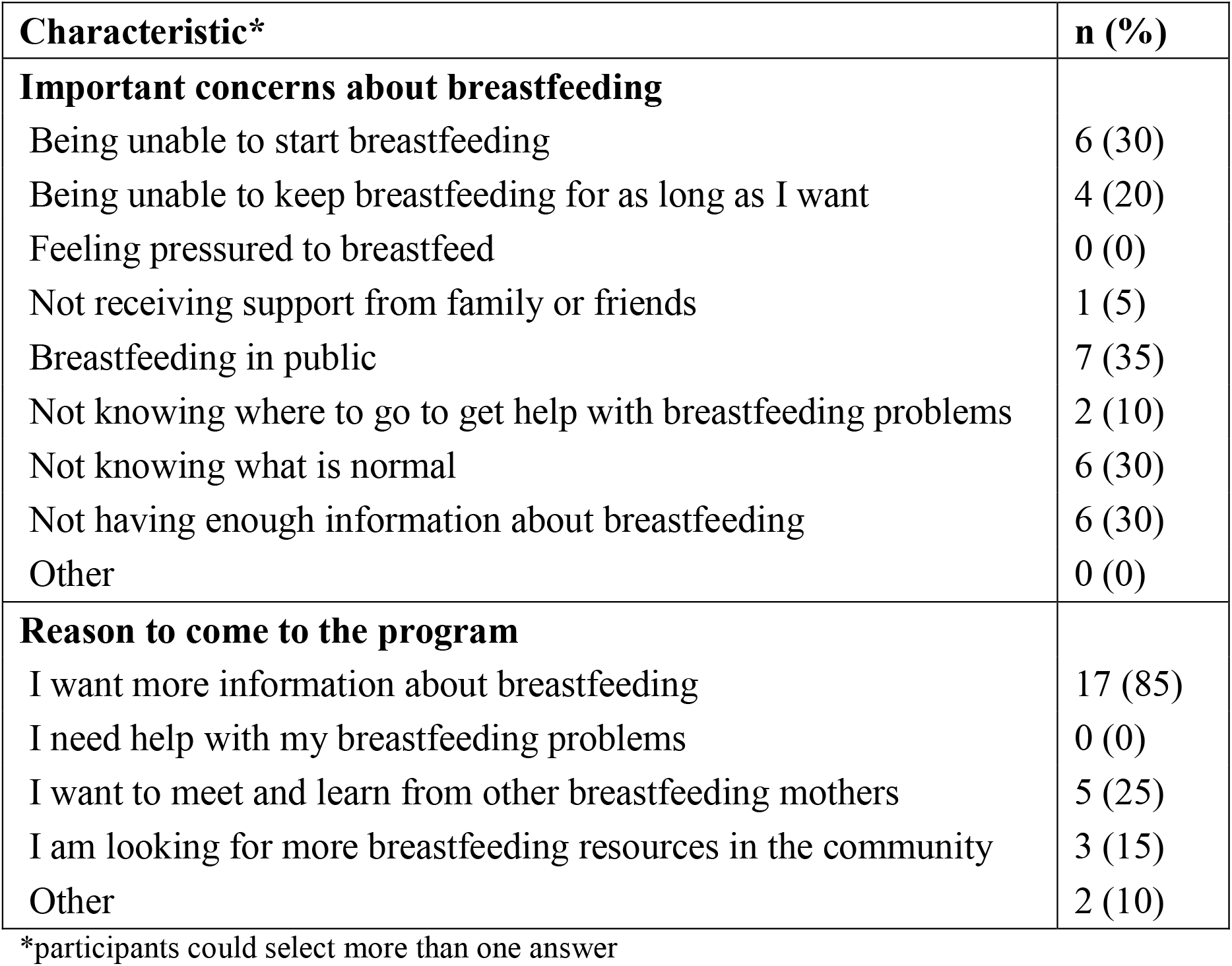
Important concerns about breastfeeding and reasons for attending the program (N=20)

Complete pre/post pairs of data were available for only 7 of the participants (attending either the prenatal class and/or four sessions of the peer support group). The overall composite BFSE-SF mean and standard deviation for pre-score was 47.7 (6.5) and for the post-score was 52.9 (SD=7), indicating an improvement overall (Table 3). There was only one participant attending the *Little Milk Miracles* mom-to-mom peer support program who provided scores for all the questions. The pre- and post-intervention score for this individual was 54 and 55, respectively. Statistical comparisons were not made due to the small sample size.

**Table 3.** Pre vs post overall BSES-SF score in all those who have complete data (N= 7)

| Descriptive statistic* | N | Pre Score | Post Score |
| --- | --- | --- | --- |
| **Overall** | 7 |  |  |
| Mean (SD), range |  | 46.7 (6.5) [39.0, 54.0] | 52.9 (7.0) [39.0, 60.0] |
| Median (IQR) |  | 50.0 (40.5, 51.5) | 54.0 (51.5, 57.0) |
| **Pregnancy Circle prenatal class only** | 5 |  |  |
| Mean (SD), range |  | 44.6 (6.5) [39.0, 53.0] | 52.6 (8.4) [39.0, 60.0] |
| Median (IQR) |  | 42.0 (39.0, 50.0) | 54.0 (51.0, 59.0) |
| **Attended both** | 1 |  |  |
| Mean (SD), range |  | 50.0 (NA) [50.0, 50.0] | 52.0 (NA) [52.0, 52.0] |
| Median (IQR) |  | 50.0 (50.0, 50.0) | 52.0 (52.0, 52.0) |

\*Due to limited sample size, cannot calculate p-value.

Scores for individual BSES-SF items before and after the BF program are presented in Figures1 and 2. Generally, the shifting of the color bars to right post-intervention indicate a higher proportion of participants that are confident (or very confident). For example, the second graph in Figure 1, “I think that I can always determine that my baby is getting enough” indicates that pre-intervention, 29% of the study population were “not at all/ not very confident”; 43% were “sometimes confident”; and 29% were “confident/very confident”. Post-intervention these proportions changed to 43% were “sometimes confident” and 57% were “confident/very confident”.

**Figure 1.**
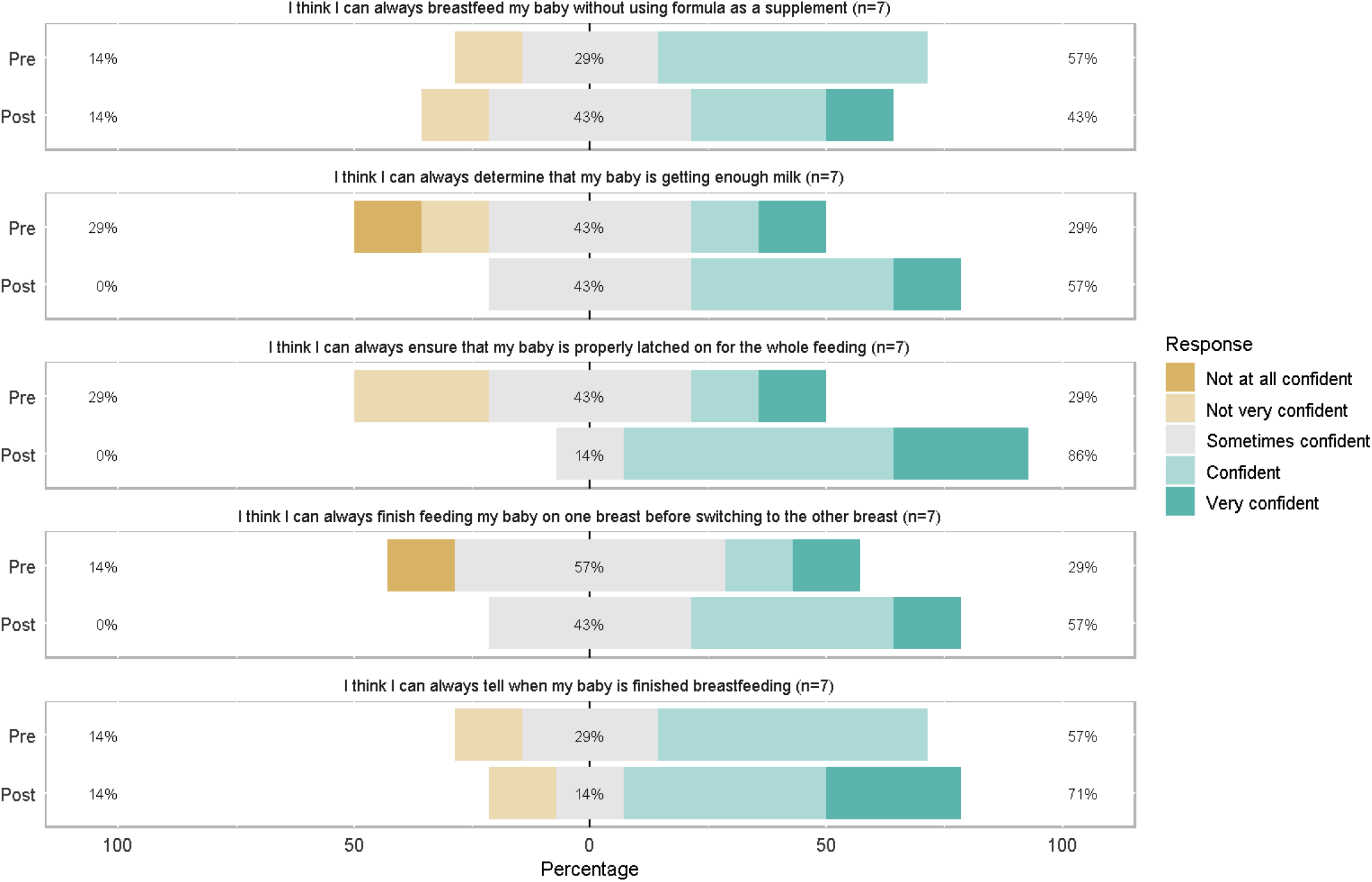
Breastfeeding Self-Efficacy Scale Items related to Knowledge.

**Figure 2.**
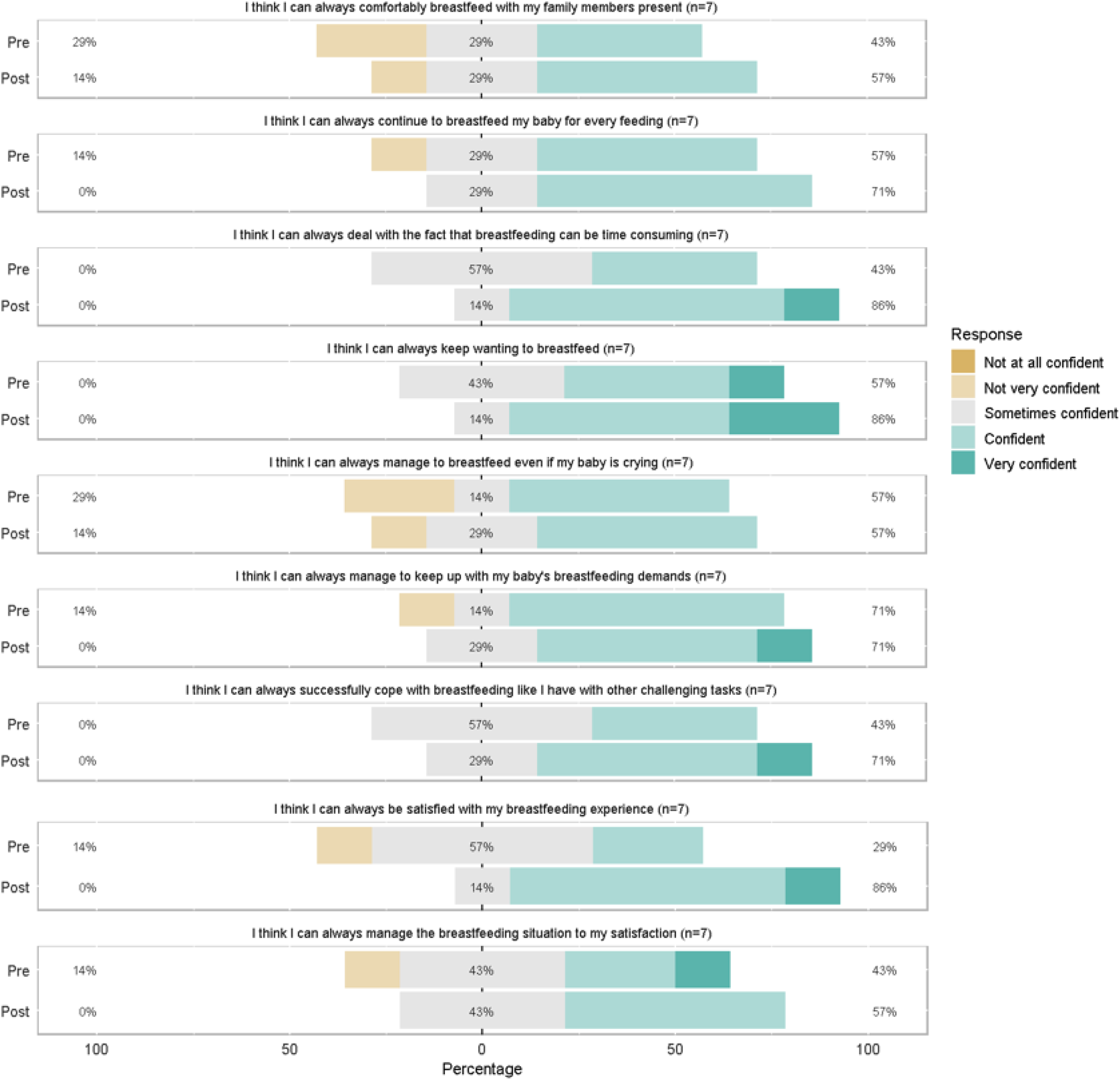
Breastfeeding Self-Efficacy Scale Items related to Coping and Satisfaction.

Data were also collected on clients up to 24 years old given this is the population the agency serves. The same analysis was done including data from these individuals (N=32) and the results were consistent with participants 19 years and younger.

## Discussion

This cohort study showed that adolescent mothers in Eastern Ontario, enrolled in a tailored youth-informed BF program, had increased BF SE. A validated tool, the BSES-SF, was used to evaluate the impact of the program which is a strength of this study. The findings are clinically relevant as it appears that participation in the BF program was associated with a change from low to high BF SE. To our knowledge, this is the only prospective study exploring BF SE of participants attending a youth-focused BF program. Previous researchers have identified a BSES-SF cutoff score of 52 or greater demonstrates high confidence. Determining BSES-SF scores is important because it predicts BF initiation, duration and exclusivity (Dennis et al., 2011; Mossman et al., 2008). The overall pre vs post comparison of composite BSES-SF scores indicates that there was an improvement in the young mothers’ SE after attending the program, indicating that the BF program improved the mothers’ knowledge, confidence and satisfaction in BF. The only item that did not demonstrate an overall improvement was BF without using formula as a supplement. This is an area that warrants further investigation as it is not known whether participants had intended to combination feed or had encountered challenges resulting in supplementation with formula.

Two strengths of this program is that it provides anticipatory guidance and timely support. Pregnant adolescents need to understand the basic mechanisms of breastfeeding (i.e. milk production is triggered by stimulation at the breast and emptying of the breast; colostrum changes to transitional milk in the first few days, and then mature milk over the first week after birth; newborns have small stomachs and therefore do not require large volumes of colostrum/milk; feedings are frequent in the early weeks after birth as breastmilk is easily digestible, etc.). By understanding “what is normal”, adolescents will be reassured that BF is proceeding as expected and perhaps they will not be tempted to supplement with formula. Hinic (2016) reported that BSES-SF scores were greater in women with previous BF experience and lowest in women who received in-hospital supplementation. Given that most adolescents are unlikely to have BF experience, anticipatory guidance and timely support received from peers and professionals is critical; our peer support program effectively provides both.

Nesbitt et al. (2012) identified, from their qualitative study of 16 adolescent mothers (aged 15-19), five influencers on continued BF including social support and knowledge. The authors suggest that professionals can engage with adolescent mothers in both the prenatal and postnatal period to assess for potential barriers to BF, to share evidence-based information, and to provide support and BF skill development. The inclusion of positive social support networks was also emphasized as an important component of BF support. It is evident that comprehensive BF support is required, particularly for populations with low rates of BF.

This study has the following limitations: Firstly, our sample was small and limited to young women in Eastern Ontario who were attending a tailored program with co-located services; it is possible that adolescents in other communities do not have access to this type of program. Although adolescents would have similar sociodemographic realities, the results may not be generalizable to other populations.

Secondly, the BF peer support program was new and as such it was challenging to recruit participants and retain them in the study for four weeks. There were only 7 complete data sets and this limited the analysis that could be completed. Adolescents are difficult to engage in services and retain as participants in research studies due to their transient nature. Further exploration of the non-respondents (post intervention) is required to determine if they had characteristics that made them inherently different than the participants who answered both surveys. This information would potentially provide valuable insight into defining the targeted support required for adolescents who are at-risk for not BF. Lastly, it is not known if increases in BSES-SF scores were associated with improved BF outcomes such as longer BF duration as this was not measured in the current study.

### Implication for Practice, Policy and Research

The challenge for policy makers and health care service organizations is to determine the ideal interventions to promote optimal population health. In the case of breastfeeding, the interventions are known. In their recent Cochrane review, McFadden et al. (2017) outlined the characteristics of effective BF support for healthy, term mother-baby dyads: predictability, scheduled, and ongoing. These characteristics enhance BF support by allowing mothers to anticipate when help will be available. Additionally, the authors emphasized that support should be tailored to the setting and the needs of the population. It is important to note that “support may be offered either by professional or lay/peer supporters, or a combination of both” and that “strategies that rely mainly on face-to-face support are more likely to succeed with women practicing exclusive breastfeeding” (McFadden et al., 2017, p. 3). These are all characteristics of our BF program. Furthermore, it is important to note that this program takes place within a one-stop, youth outreach agency which offers a full range of programs and services such as a medical care, as well as multiple social and educational programs such as prenatal classes, parenting classes and the new BF program (St. Mary’s Home, 2020). Childcare and transportation assistance is available which can significantly facilitate attendance and participation of at-risk adolescents. These attributes likely contributed to the success of this program.

The US Preventive Services Task Force, in their updated evidence report and systematic review on primary care intervention to support BF, confirmed that BF support interventions are associated with an increase in the rates of any BF as well as exclusive BF (Patnode, et al., 2016). Recommendations from this review emphasize the importance of providing interventions during pregnancy and after birth including: promoting the benefits of BF, providing practical advice and direct support on how to breastfeed, and providing psychological support. In our tailored program, the prenatal BF class addresses the first recommendation and the peer support group accomplishes all of the recommendations.

Sipsma and colleagues (2015), in their systematic review of BF promotion among adolescent mothers, identified that a combination of education and counseling provided by a lactation consultant–peer counselor team significantly improved both BF initiation and duration. This is consistent with the systematic reviews of breastfeeding support among adults (Lumbiganon, Martis, Laopaiboon, Festin, Ho, & Hakimi, 2012; McFadden et al., 2017; Patnode, Henninger, Senger, Perdue, Whitlock, 2016; Renfrew, McCormick, Wade, Quinn, & Dowswell, 2012). The findings from our cohort study are promising as they indicated an improved SE of adolescent mothers attending the tailored evidence-based BF program (which included antenatal education and peer support). These types of programs need to be adequately funded if the suboptimal BF rates are to be improved.

## Conclusion

Prenatal education and peer support adapted to the needs of adolescents appears to be associated with increased BSES-SF scores. Identifying mothers with low BF SE can enable health care professionals to implement targeted interventions with the goal of achieving longer BF rates in this at-risk population. Given the promising results of this cohort study, further research with a larger cohort is warranted to determine if participation in the youth-informed BF program is associated with improved outcomes such as increased rate of BF intention, initiation and duration in adolescents.

## Data Availability

Available upon request.

## Funding Acknowledgement

The Innovative Youth-Informed Breastfeeding Program was developed and delivered with funds provided by the Government of Ontario and the support of the Best Start Resource Centre at Health Nexus.

## Acknowledgements

We would like to acknowledge the contributions of the following individuals in the development of the Innovative Youth-Informed Breastfeeding Program: Jill Behn, Beverley Croft, Cathryn Fortier, Julie Gagnier, Maria Gareau, Marianne Gervais, Kim Ledoux, Nancy MacNider, and Nicole Olive.

## References

Apostolakis-Kyrus, K., Valentine, C., & DeFranco, E. (2013). Factors associated with breast-feeding initiation in adolescent mothers. The Journal of Pediatrics, 163(5), 1489–1494.

Best Start Resource Centre (2015a). Populations with lower rates of breastfeeding: A summary of findings. Toronto, ON: Author.

Best Start Resource Centre (2015b). Fact sheet #4.Breastfeeding peer support programs: An effective strategy to reach and support populations with lower rates of breastfeeding. Retrieved from http://www.beststart.org/resources/breastfeeding/B18-E_Breastfeeding_factsheet_4_rev.pdf

Best Start Resource Centre (2015c). Developing and sustaining peer support programs. Toronto, ON: Author.Retrieved from http://www.beststart.org/resources/breastfeeding/B10_BF_Peer_Support_Programs_ENG_final.pdf

Best Start Resource Centre (2017). Breastfeeding community projects. 2015 - Round 2 breast-feeding community projects. Retrieved from http://en.beststart.org/services/partnerships-and-projects/breastfeeding-community-project

Dennis, C.-L., Heaman, M., & Mossman, M. (2011). Psychometric testing of the breastfeeding self-efficacy scale-short form among adolescents. Journal of Adolescent Health, 49, 265–271.

Durham Region Health Department (2015, June). Infant feeding surveillance system. Focused report on infant feeding among adolescent mothers. Durham, ON: Author. Retrieved from http://www.durham.ca/departments/health/health_statistics/IFSSadolescentMothers.pdf

Fleming, N. A., Tu, X., & Black, A. Y. (2012). Improved obstetrical outcomes for adolescents in a community-based outreach program: A matched cohort study. Journal of Obstetrics and Gynaecology of Canada, 34(12), 1134–1140.

Fleming, N., Ng, N., Osborne, C., Biederman, S., Ill, A. S., Dy, J., … Walker, M. (2013). Adolescent pregnancy outcomes in the province of Ontario: A cohort study. Journal of Obstetrics and Gynaecology of Canada, 35(3), 234–245.

Gionet, L. (2013, November). Health at a glance-Breastfeeding trends in Canada. Retrieved from Statistics Canada: http://www.statcan.gc.ca/pub/82-624-x/2013001/article/11879-eng.pdf

Health Canada. (2015, August 18). Nutrition for healthy term infants: Recommendations from birth to six months. Retrieved October 15, 2016, from Health Canada: http://www.hc-sc.gc.ca/fn-an/nutrition/infant-nourisson/recom/index-eng.php#a3

Hinic, K. (2016). Predictors of breastfeeding confidence in the early postpartum period. Journal of Obstetric, Gynecologic and Neonatal Nursing, 45(5), 649–660. doi: 10.1016/j.jogn.2016.04.010

Leclair, E., Robert, N., Sprague, A. E., & Fleming, N. (2015). Factors associated with breast-feeding initiation in adolescent pregnancies: A cohort study. Journal of Pediatric and Adolescent Gynecology, 28(6), 516–521.

Lumbiganon, P., Martis, R., Laopaiboon, M., Festin, M. R., Ho, J. J., & Hakimi, M. (2012). Antenatal breastfeeding education for increasing breastfeeding duration. Cochrane Database of Systematic Reviews, Issue 9. Art. No.: CD006425. doi:10.1002/14651858.CD006425.pub3.

McFadden, A., Gavine, A., Renfrew, M. J., Wade, A., Buchanan, P., Taylor, J. L., … MacGillivray, S. (2017). Support for healthy breastfeeding mothers with healthy terms babies. Cochrane Database of Systematic Reviews 2017, Issue 2. Art. No.: CD001141. DOI: 10.1002/14651858.CD001141.pub5.

Mossman, M., Heaman, M., Dennis, C.-L., & Morris, M. (2008). The influence of adolescent mothers’ breastfeeding confidence and attitudes on breastfeeding initiation and duration. Journal of Human Lactation, 24(3), 268–277.

Nesbitt, S. A., Campbell, K. A., Jack, S. M., Robinson, H., Piehl, K., & Bogdan, J. C. (2012). Canadian adolescent mothers’ perceptions of influences on breastfeeding decisions: a qualitative descriptive study. BMC Pregnancy and Childbirth, 12.

Ontario Ministry of Health and Long Term Care (2014). Transforming Ontario’s health care system: 2013 Minister’s medal winners.Individual champion recipient: Dr. Nathalie Fleming, Young Parent Outreach Centre. Retrieved from http://www.health.gov.on.ca/en/pro/programs/transformation/minister_medal_2013.aspx

Ottawa Public Health (2013). BFI Expo 2013. Public health panel-Ottawa Public Health. Retrieved from http://www.bfiontario.ca/wp-content/uploads/2013/02/BFI-Expo-2013-Public-Health-Panel-Ottawa-Public-Health.pdf

Patnode, C. D., Henninger, M. L., Senger, C. A., Perdue, L. A., & Whitlock, E. P. (2016). Primary care intervention to support breastfeeding. Updated evidence report and systematic review for the US Preventive Services Task Force. Journal of the American Medical Association, 316(16), 1694–1705.

R Core Team (2017). R: A language and environment for statistical computing. R Foundation for Statistical Computing, Vienna, Austria. https://www.r-project.org/

Renfrew, M. J., McCormick, F. M., Wade, A., Quinn, B., & Dowswell, T. (2012). Support for healthy breastfeeding mothers with healthy term babies. Cochrane Database of Systematic Reviews, Issue 5. Art. No.: CD001141. doi:10.1002/14651858.CD001141.pub4.

aSt. Mary’s Home (2020). About us. Retrieved from http://stmaryshome.com

Sipsma, H. L., Jones, K. L. & Cole-Lewis, H. (2015). Breastfeeding among adolescent mothers: A systematic review of interventions from high-income countries. Journal of Human Lactation, 31(2), 221–229.

Sipsma, H., Magriples, U., Divney, A., Gordon, D., Gabzdyl, E., & Kershaw, T. (2013). Breastfeeding behavior among adolescents: Initiation, duration, and exclusivity. Journal of Adolescent Health, 53(3), 394–400.

World Health Organization. (2020). Maternal Newborn Child and Adolescent Health: Breastfeeding. Retrieved from http://www.who.int/maternal_child_adolescent/topics/newborn/nutrition/breastfeeding/en Accessed February 16, 2018.

